# Virologic features of SARS-CoV-2 infection in children

**DOI:** 10.1101/2021.05.30.21258086

**Authors:** Lael M. Yonker, Julie Boucau, James Regan, Manish C. Choudhary, Madeleine D. Burns, Nicola Young, Eva J. Farkas, Jameson P. Davis, Peter P. Moschovis, T. Bernard Kinane, Alessio Fasano, Anne M. Neilan, Jonathan Z. Li, Amy K. Barczak

## Abstract

**Background:** Data on pediatric COVID-19 has lagged behind adults throughout the pandemic. An understanding of SARS-CoV-2 viral dynamics in children would enable data-driven public health guidance.

**Methods:** Respiratory swabs were collected from children with COVID-19. Viral load was quantified by RT-PCR; viral culture was assessed by direct observation of cytopathic effects and semiquantitative viral titers. Correlations with age, symptom duration, and disease severity were analyzed. SARS-CoV-2 whole genome sequences were compared with contemporaneous sequences.

**Results:** 110 children with COVID-19 (median age 10 years, range 2 weeks-21 years) were included in this study. Age did not impact SARS-CoV-2 viral load. Children were most infectious within the first five days of illness, and severe disease did not correlate with increased viral loads. Pediatric SARS-CoV-2 sequences were representative of those in the community and novel variants were identified.

**Conclusions:** Symptomatic and asymptomatic children can carry high quantities of live, replicating SARS-CoV-2, creating a potential reservoir for transmission and evolution of genetic variants. As guidance around social distancing and masking evolves following vaccine uptake in older populations, a clear understanding of SARS-CoV-2 infection dynamics in children is critical for rational development of public health policies and vaccination strategies to mitigate the impact of COVID-19.

## Background

Since the SARS-CoV-2 virus ignited the COVID-19 global pandemic, the impact of the virus on children and the role that children play in this pandemic has been understudied. Initially, epidemiology reports suggested that children may have been relatively spared from infection, however, as COVID-19 testing became more available, it has been increasingly recognized that children can be infected with SARS-CoV-2 at rates comparable to adults [1, 2]. To date, over 4.1 million children in the Unites States have been reported as testing positive for COVID-19 [3]. Since the winter of 2020-2021, children under 19 years of age have represented one of the age groups with the highest rates of infection [4], which likely reflects a combination of increased number of infections among children plus increased vaccination rates amongst adults. Most children generally have milder symptoms when infected with SARS-CoV-2 [5], although a small subset of individuals develop severe disease. In the US, over 16,000 children have been hospitalized for acute COVID-19 with over 300 deaths reported[3]. A baseline understanding of the viral characteristics of SARS-CoV-2 infection in children is a necessary prerequisite to understanding the pathogenesis of severe presentations of COVID-19 [6].

At a population perspective, the role that children play in viral transmission remains poorly understood. Epidemiologic studies suggest that children exhibit lower transmission rates than adults [7], however, these findings are potentially confounded by higher rates of asymptomatic or pauci-symptomatic infection in children, increased social isolation by children early in the pandemic, and reduced COVID-19 testing in children. To date, one small study demonstrated that live virus can be cultured from children [8]. However, the types of systematic studies that have informed our understanding of the viral dynamics of SARS-CoV-2 in adult populations [9-11] have not similarly been carried out in children. As vaccination has become available for adults and adolescents in many places in the world and our understanding of transmission dynamics have evolved, masking and distancing policies are being relaxed[12]. Policy changes have necessarily been made despite the paucity of data providing insight into the role that pediatric disease might play in ongoing transmission. As viral variants that enhance the potential for transmission and/or reduce vaccine efficacy emerge [13-15], the importance of identifying potential reservoirs of viral replication and transmission has been brought into the spotlight. Defining the virologic features of SARS-CoV-2 infection in children and the potential for children to transmit virus will facilitate rational public health decision-making for pediatric populations.

In this work, we sought to define fundamental virologic features of SARS-CoV-2 in a pediatric population across a range of disease severity. We analyzed respiratory swabs from children presenting to urgent care clinics or the hospital with symptomatic and asymptomatic COVID-19 infection. Clinical factors, such as age, COVID-19 risk factors, and disease severity were compared with viral features including SARS-CoV-2 viral load, isolation of replication-competent virus, and whole viral sequencing. Our data indicate that age, from infancy through adulthood, is not a predictor of viral infection dynamics, and that children of all ages can have high SARS-CoV-2 viral loads of replication-competent virus, including variants, displaying comparable dynamics to those seen in adults.

## Methods

### Sample collection

Infants, children and adolescents ≤21 years of age presenting to Massachusetts General Hospital urgent care clinics or the hospital with either symptoms concerning for or known exposure to COVID-19 (4/2020-4/2021) were prospectively offered enrollment in the Institutional Review Board-approved MGH Pediatric COVID-19 Biorepository (IRB # 2020P000955) [16]. After informed consent, and assent when appropriate, was obtained verbally, a research-designated swab of the nasopharynx, oropharynx and/or anterior nares was obtained and placed in phosphate buffered saline. Samples were aliquoted and stored at -80°C. Samples from patients who tested positive for COVID-19 on clinical SARS-CoV-2 RT-PCR testing were analyzed. Nasal samples from adults hospitalized with acute COVID-19 [10] (4/2020-8/2020; enrolled in Institutional Review Board-approved MGH COVID-19 Biorepository, IRB # 2020P000804) with duration of symptoms equal to the hospitalized pediatric cohort were selected for comparative studies.

### Clinical data collection

Demographic and clinical factors were recorded through a combination of manual chart reviews and data extraction from electronic health records (EHR), then collected in a REDCap database [17] through the Partners Electronic Health Record Registry of Pediatric COVID-19 Disease (IRB # 2020P003588). Trained reviewers collected demographics, SARS-CoV-2 risk factors, comorbid conditions, medications, COVID-19 related symptoms, and laboratory tests. Outcome of initial presentation to care, admission status, and complications of COVID-19 disease were also extracted by manual review.

### SARS-CoV-2 viral load quantification

Virions were pelleted from anterior nasal, oropharyngeal, and nasopharyngeal swab fluids by centrifugation at approximately 21,000 x g for 2 hours at 4°C. RNA was extracted using Trizol-LS (Thermofisher) according to the manufacturer’s instructions. RNA was then concentrated by isopropanol precipitation, and SARS-CoV-2 RNA was quantified using the CDC N1 primers and probe [https://www.cdc.gov/coronavirus/2019-ncov/lab/rt-pcr-panel-primer-probes.html] as previously described [10]. As there was no significant difference in viral load from respiratory secretions obtained from the anterior nares, nasopharynx or oropharynx of participants (**Supplemental Figure 1**), samples were analyzed together, regardless of collection site.

### Viral Culture

Vero-E6 cells (ATCC) were maintained in D10+ media [Dulbecco’s modified Eagle’s media (DMEM) (Corning) supplemented with HEPES (Corning), 1X Penicillin/Streptomycin (Corning), 1X Glutamine (Glutamax, ThermoFisher Scientific) and 10% Fetal Bovine serum (FBS) (Sigma)] in a humidified incubator at 37°C in 5% CO2. Vero-E6 cells were passaged every 3-4 days, detached using Trypsin-EDTA (Fisher Scientific) and seeded at 150,000 cells per wells in 24 well plates for culture experiments and 20,000 cells per well in 96w plates the day before inoculation for median tissue culture infectious dose (TCID50) experiments.

After thawing, each specimen was filtered through a Spin-X 0.45µm filter (Corning) at 10,000 x g for 5min. 50µL of the supernatant was then diluted in 450µL of D^+^ media [DMEM supplemented with HEPES, 1X Penicillin/Streptomycin and 1X Glutamine]. The viral culture experiments were performed as previously reported [18] with the following modifications: 100µL of the solution was used to inoculate wells in a 24 well plate and 1mL of D_2_^+^ media [D+ media with 2% FBS] was added to each well after 1 hour of incubation. The plates were then placed in a 5% CO2 incubator at 37°C. For TCID50 measurements conducted in parallel, 25µL of the Spin-X flow-through was used to inoculate Vero-E6 cells in a 96 well plate in the presence of 5µg/mL of polybrene (Santa Cruz Biotechnology) using 5-fold dilutions (5^-1^ to 5^-6^) and 4 repeats for each sample. The plates were centrifuged for 1 hour at 2,000 x g at 37°C before being placed in a 5% CO2 incubator at 37°C. The SARS-CoV-2 isolate USA-WA1/2020 strain (BEI Resources) was used as a positive control for CPE in both culture and TCID50 experiments.

Viral culture and TCID50 plates were observed at 3- and 6-days post-infection with a light microscope and wells showing CPE were counted. The TCID50 titers were calculated using the Spearman-Karber method. For the culture plates, the supernatant of the wells displaying CPE was harvested 10-14 days post-infection and RNA was isolated using a QIAamp Viral RNA Mini kit (QIAGEN) for confirmation of the viral sequence.

### SARS-CoV-2 sequencing

cDNA synthesis was performed using Superscript IV reverse transcriptase (Invitrogen). Whole viral amplification was performed with the Artic protocol using multiplexed primer pools designed with Primal Scheme generating 400-bp tiling amplicons [19, 20]. PCR products were pooled and Illumina library construction was performed using the Nextera XT Library Prep Kit (Illumina). The comparison dataset included 183 representative contemporaneous SARS-CoV-2 genomes from Massachusetts present in GISAID to assess for local clustering. Nucleotide sequence alignment was performed with MAFFT (multiple alignment using fast Fourier transform) [21]. Best-fit nucleotide substitution GTR+G+I was used for the datasets using model selection in IQ-Tree followed by maximum likelihood phylogenetic tree construction using IQ-Tree web server with 1000-bootstrap replicates [22].

### Analysis

All statistical analyses were performed using parametric comparisons in GraphPad Prism (Version 9.1.1), including Pearson correlation, ANOVA with multiple comparisons, and unpaired t test.

## Results

### Clinical cohort

One-hundred-ten children diagnosed with COVID-19 with a mean age of 10 years (range 0-21 years) were included in the study (**Table 1**). There were slightly more boys (56%) than girls (44%) with SARS-CoV-2 infection included in our analyses. One third of the participants were White (33%), 10% African American/Black, and 4% were Asian; one third (38%) reported their ethnicity as Hispanic. Past medical history in children is reported in **Supplemental Table 1**. Thirty children were asymptomatic but were identified as having COVID-19: twenty-six children (27%) presented to urgent care/COVID-19 testing sites because of a COVID-19 exposure, while four children (4%) were identified on routine screening during hospital admission. Eight (7%) presented with COVID-19 symptoms but had no known COVID-19 contact. The majority of participants with COVID-19 did not require hospitalization (72 children, 65%). Thirty-six children (33%) were hospitalized with COVID-19, although only 18 children (16%) required supplemental oxygen and/or invasive or non-invasive respiratory support (referred to as “moderate/severe COVID-19”).

**Table 1:** Participant Demographics, past medical history, reason for presenting for SARS-CoV-2 RT-PCR testing, and disease severity of children with COVID-19 (n=110).

|  | **COVID+ children (N=110)** |
| --- | --- |
| *Age, average (max, min)* | 10 (0,21) |
| *Sex, n (%)* |  |
| Male | 62 (56) |
| Female | 48 (44) |
| *Race, n (%)* |  |
| White | 36 (33) |
| Black | 11 (10) |
| Asian | 4 (4) |
| Other | 45 (41) |
| *Ethnicity, n (%)* |  |
| Hispanic | 42 (38) |
| *Past Medical History, n (%)* |  |
| Obesity | 28 (25) |
| Asthma | 11 (10) |
| Other | 39 (35) |
| *Reason for presenting for COVID-19 testing, n (%)* |  |
| Asymptomatic, exposure | 30 (27) |
| Symptomatic, exposure | 72 (65) |
| Symptomatic, no exposure | 8 (7) |
| *COVID-19 Severity, n (%)* |  |
| Hospitalized | 36 (33) |
| Supplemental oxygen | 18 (16) |
| Outpatient | 75 (68) |

### Age did not impact SARS-CoV-2 viral load or recovery of replication competent virus

Age is a well-established risk factor for developing severe COVID-19. Accordingly, asymptomatic patients were significantly younger than patients with mild disease, and pediatric patients who were hospitalized with hypoxemia were significantly older than asymptomatic children or children with mild disease (**Figure 1A**). However, viral load was not increased in more severe disease: asymptomatic children and children with mild disease displayed significantly higher viral loads than adults hospitalized with COVID-19 with comparable duration of symptoms (less than 10 days) (**Figure 1B**). However, there were no differences in viral load between pediatric patients hospitalized with moderate/severe disease and hospitalized adults of similar duration of illness (**Figure 1B**) (Adult demographics are detailed in **Supplemental Table 2**).

**Figure 1:**
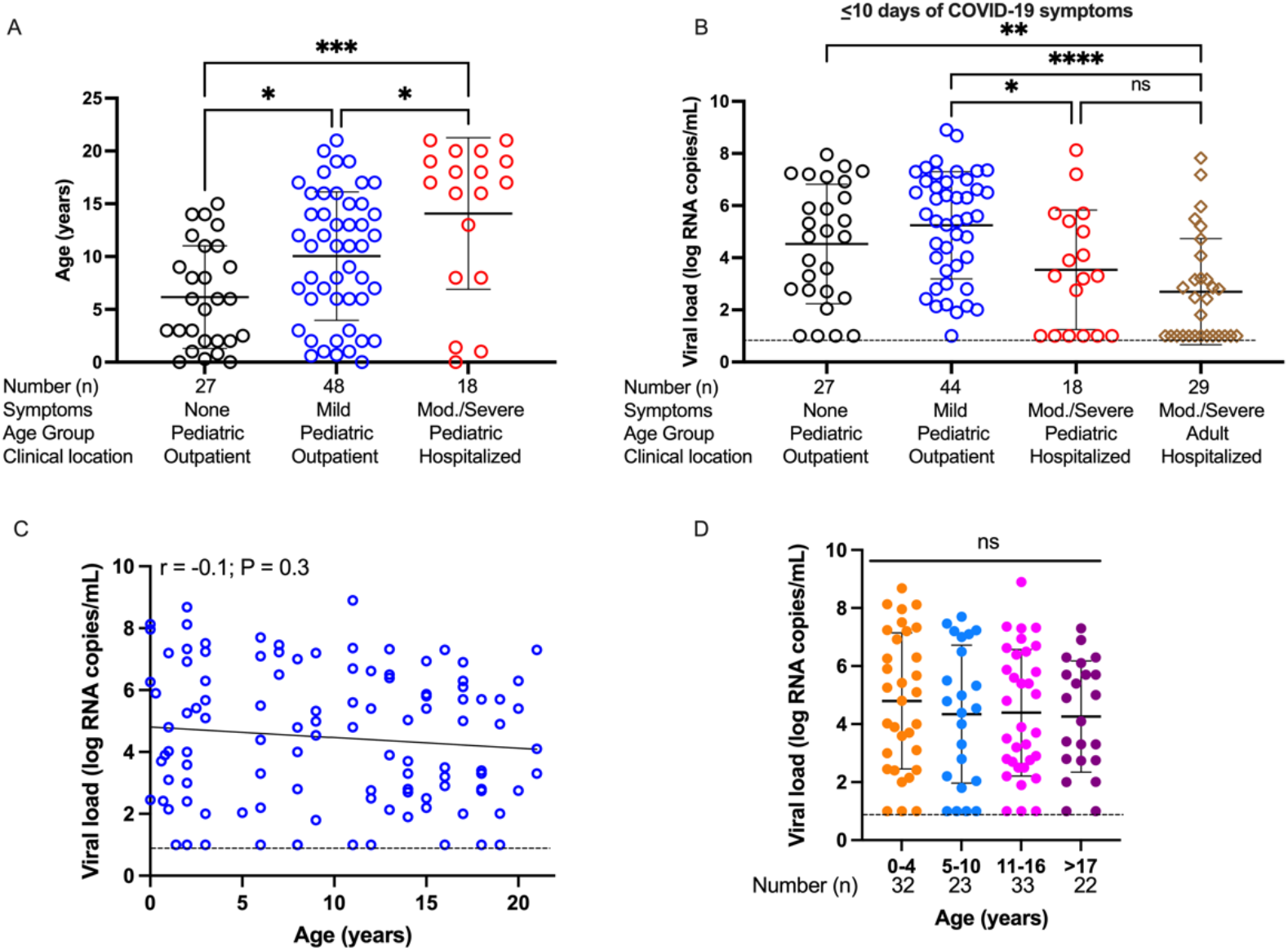
COVID-19 disease severity and SARS-CoV-2 viral load across age groups. **A**. Age of pediatric patients with SARS-CoV-2 infection, stratified by disease severity: asymptomatic (n=27), mild disease, outpatient (n=48), moderate/severe COVID-19, hospitalized (n=18). Analyzed by ordinary one-way ANOVA. **B**. SARS-CoV-2 viral load was quantified across a range of disease severities. Patients presenting with ≤10 days of symptoms were compared, including asymptomatic pediatric outpatients (n=27), mildly symptomatic pediatric outpatients (n=44), moderate/severe pediatric hospitalized patients with oxygen requirement (n=18), and moderate/severe adult hospitalized patients (n=29). Analyzed by ordinary one-way ANOVA. **C**. Viral load for each specimen (n=110) was determined by qPCR, plotted against participant age and analyzed by Pearson correlation. **D**. Viral load levels reported by school age group: 0-4 years old (yo)– infant through pre-school (n=32), 5-10yo – elementary school (n=23), 11-16yo – middle school (n=33), 17yo and over – high school and higher education (n=22). Analyzed by ordinary one-way ANOVA. Dotted lines depict limit of detection. * P<0.05, *** P<0.001, **** P< 0.0001, ns = not significant

The age of each infected child was analyzed to determine whether age impacted viral load. There was no significant correlation of age with viral load (**Figure 1C**), nor were there significant differences between ages when grouped by school levels: 0-4 years (infant through pre-school), 5-10 years (elementary school), 11-16 years (middle school), 17 and older (high school and higher education) (ANOVA, P = 0.12) (**Figure 1D**). Thus, a child’s age did not appear to impact viral load: all children, from 2 weeks through 21 years of age, were equally capable of carrying a high viral load.

As SARS-CoV-2 RNA detection by RT-PCR does not specify whether replication-competent virus is being shed, we next sought to ascertain risk factors for shedding live virus by performing viral culture assays for recoverable SARS-CoV-2 in parallel with viral load testing. From the 110 participants, we collected 126 samples; live virus was cultured from 33 samples coming from 32 participants. Of note, eight of these children with culturable SARS-CoV-2 were asymptomatic. Higher viral load was significantly predictive of shedding of live virus (t test, P < 0.0001) (**Figure 2A**). Consistent with the results for viral load, age was not correlated with viral culture results; virus was recovered from children ages 1 month through 21 years (**Figure 2B**). Semi-quantitative assessment of the amount of virus shed by an individual participant was assessed by median tissue culture infectious dose (TCID50). TCID50 for culture-positive specimens correlated strongly with viral load (Pearson correlation r = 0.7, P < 0.0001) (**Figure 2C**) but did not correlate with age across all pediatric participants (**Figure 2D**).

**Figure 2:**
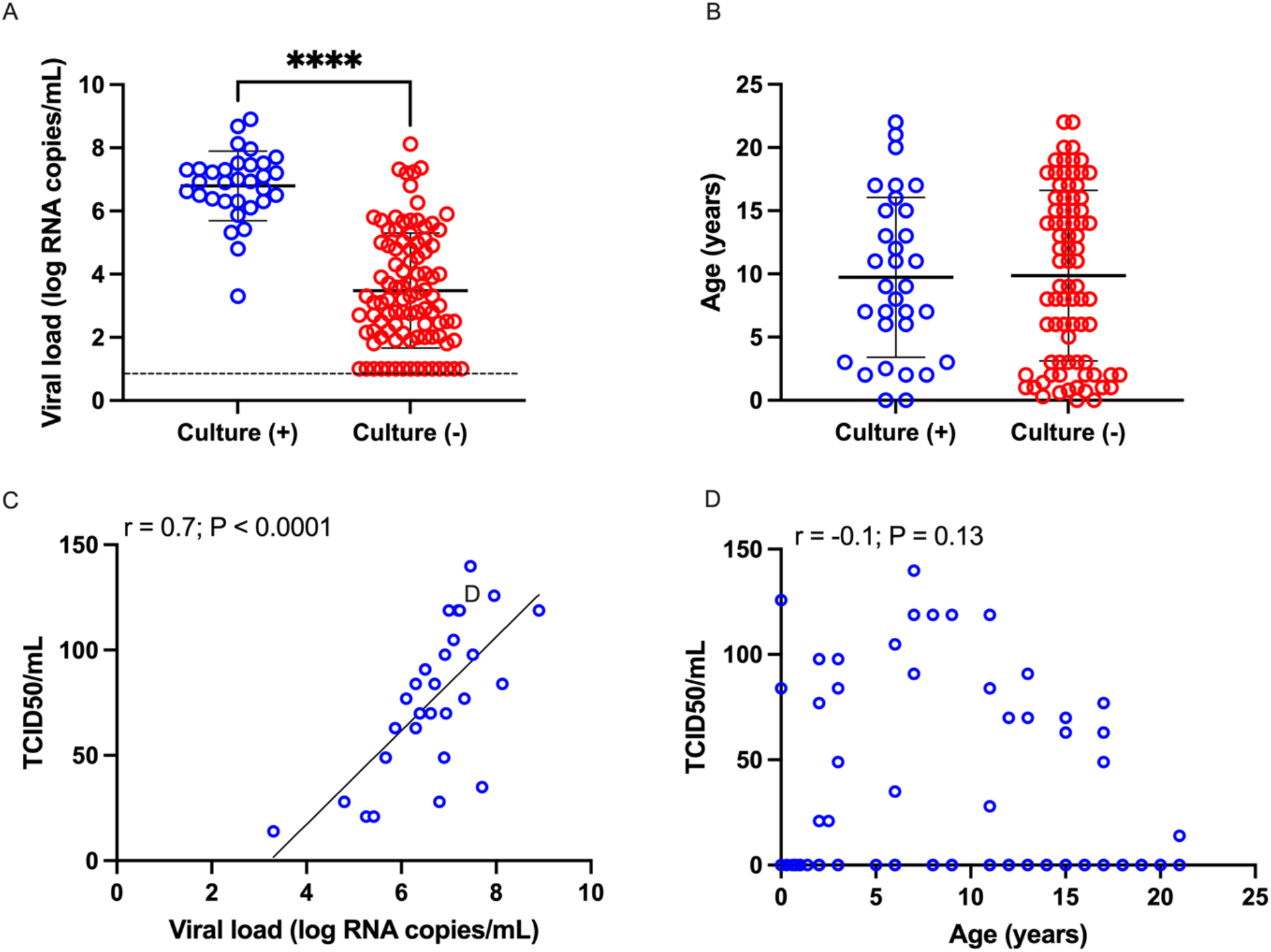
SARS-CoV-2 culture results across age groups and viral load. **A-B**. Samples with observable CPE (culture +) (n=31) or without observable CPE (culture -) (n=95) plotted against viral load (**A**) and participant age (**B**) and compared using t test. **C-D**. Semiquantitative viral titer expressed as TCID50/mL for culture positive samples plotted against corresponding viral load (**C**) or participant age (**D**), Analyzed using Pearson correlation. Dotted line depicts limit of detection. **** P < 0.0001

### Children with COVID-19 were most infectious within first five days of illness

To define the likely period of infectiousness in our pediatric population, we analyzed viral load, culturability, and TCID50 in comparison with duration of symptoms. Of note, duration of symptoms does not necessarily indicate duration of infection, as time infection was acquired cannot be confirmed. Consistent with prior reports in adults [9], viral loads in children were the highest earliest in the course of illness and declined over time after symptom onset (Pearson, r = -0.4, P <0.001) (**Figure 3A**). Viral load was highest in the first two days of symptoms, with significant decrease after 5 days of symptoms and further decline after 10 days of symptoms (P < 0.0001) (**Figure 3B**). Analysis of pediatric viral culture results demonstrated that children tested early after symptom onset were more likely to shed replication competent virus (P = 0.004) (**Figure 3C)**. Correspondingly, semi-quantitative assessment of infectious viral shedding in children showed that the TCID50 was higher early after symptom onset and decreased over time. When grouped by days of symptoms, children in days 0-2 of their symptoms had the highest infectivity, while children with greater than six days of illness shed less virus (P = 0.004) (**Figure 3D**).

**Figure 3:**
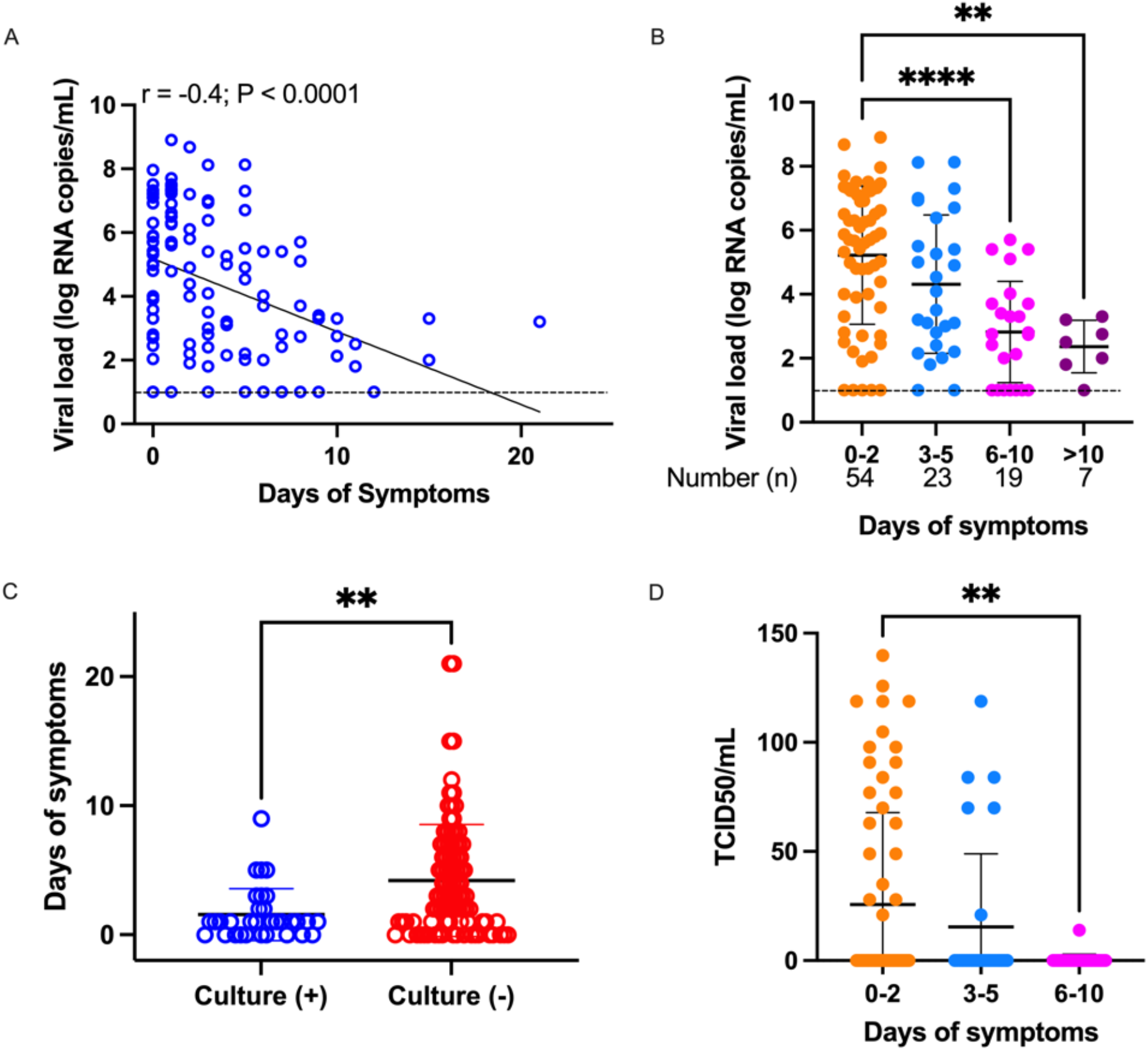
Culture positivity and duration of symptoms. **A**. Viral load for each specimen was determined by qPCR and plotted against the duration of symptoms (in days). Analysis by Pearson correlation. **B**. Viral load reported by binned duration of symptoms. Ordinary one-way ANOVA used for analysis. **C**. Duration of symptoms for samples with observable CPE (culture +, n=29) and without observable CPE (culture -, n=85). Analysis by t test. **D**. Semiquantitative viral titer reported by binned duration of symptoms, analyzed by ordinary one-way ANOVA. Dotted lines depict limit of detection. ** P < 0.01, **** P < 0.0001

We then sought to assess whether COVID-19 severity impacted the relationship between viral load and age in pediatric cohorts of varying severity: asymptomatic, mildly symptomatic, and moderate/severe pediatric COVID-19 patients. None of these COVID-19 severity groups revealed any correlation of age with viral load (**Figure 4A**). Further, there were no differences in viral clearance over the duration of illness, not only when comparing mild pediatric COVID-19 with moderate/severe COVID-19, but also when comparing pediatric COVID-19 with adults hospitalized with COVID-19 (**Figure 4B**). Duration of symptoms did not affect the finding that viral load does not correlate with age of the infected child (**Supplemental Figure 2**).

**Figure 4:**
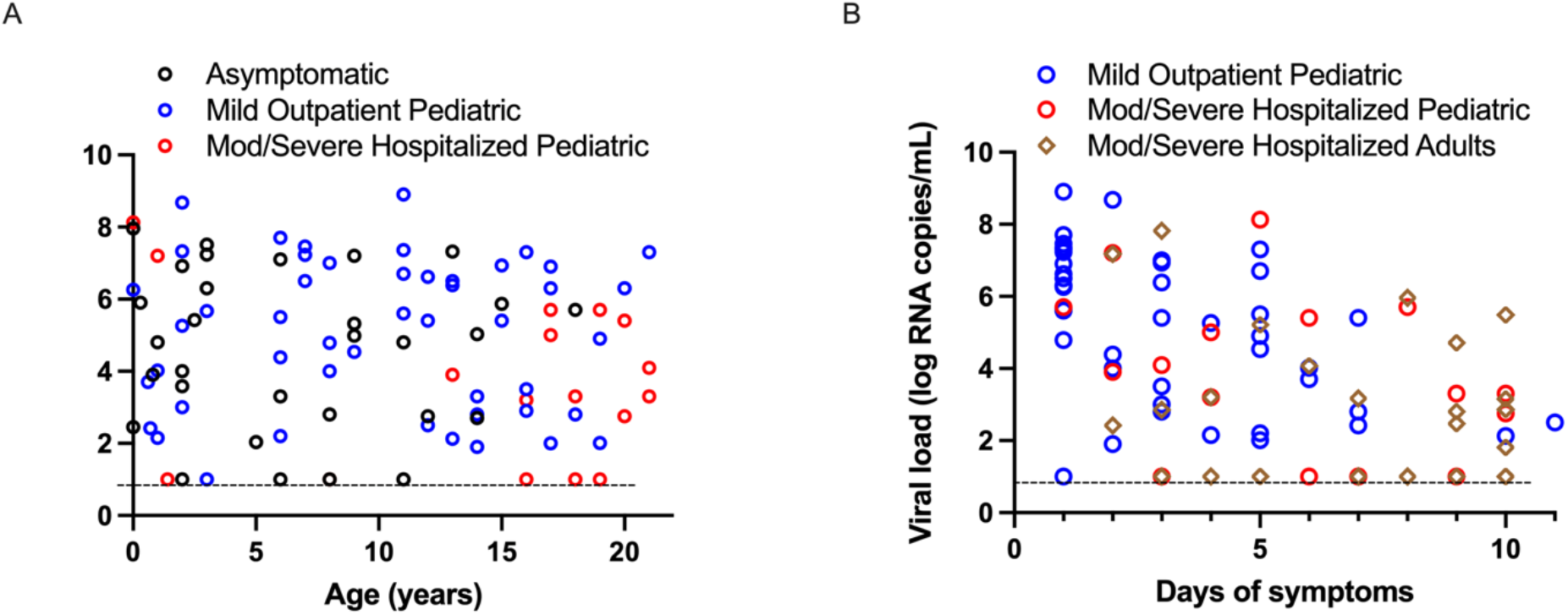
Correlation of viral load with age and duration of illness based on disease severity. **A**. Correlation of viral load and age, stratified by asymptomatic (n=30), mild outpatient (n=48) and moderate/severe hospitalized (n=18) cohorts. **B**. Viral load of hospitalized adult (n=29), hospitalized pediatric participants requiring respiratory support (n=18) and pediatric outpatients with mild disease (n=48), plotted against duration of symptoms. Dotted lines depict limit of detection.

### Pediatric SARS-CoV-2 sequences were representative of those found in the community

We successfully performed whole-viral sequencing of 57 respiratory samples from 54 children. Phylogenetic analysis of these pediatric sequences with contemporaneous Massachusetts sequences from GISAID showed that they were representative of the spectrum of sequences found in the community (**Figure 5**). Notable variants identified in the pediatric samples included four Alpha (B.1.1.7) and three lota (B.1.526.2) variants. To validate our culture results on a subset of culture-positive samples, we sequenced virus isolated from the supernatant from 8 positive samples. Sequences from the supernatant and respiratory specimens were identical in 7 cases and demonstrated only 1 nucleotide change in the last case.

**Figure 5.**
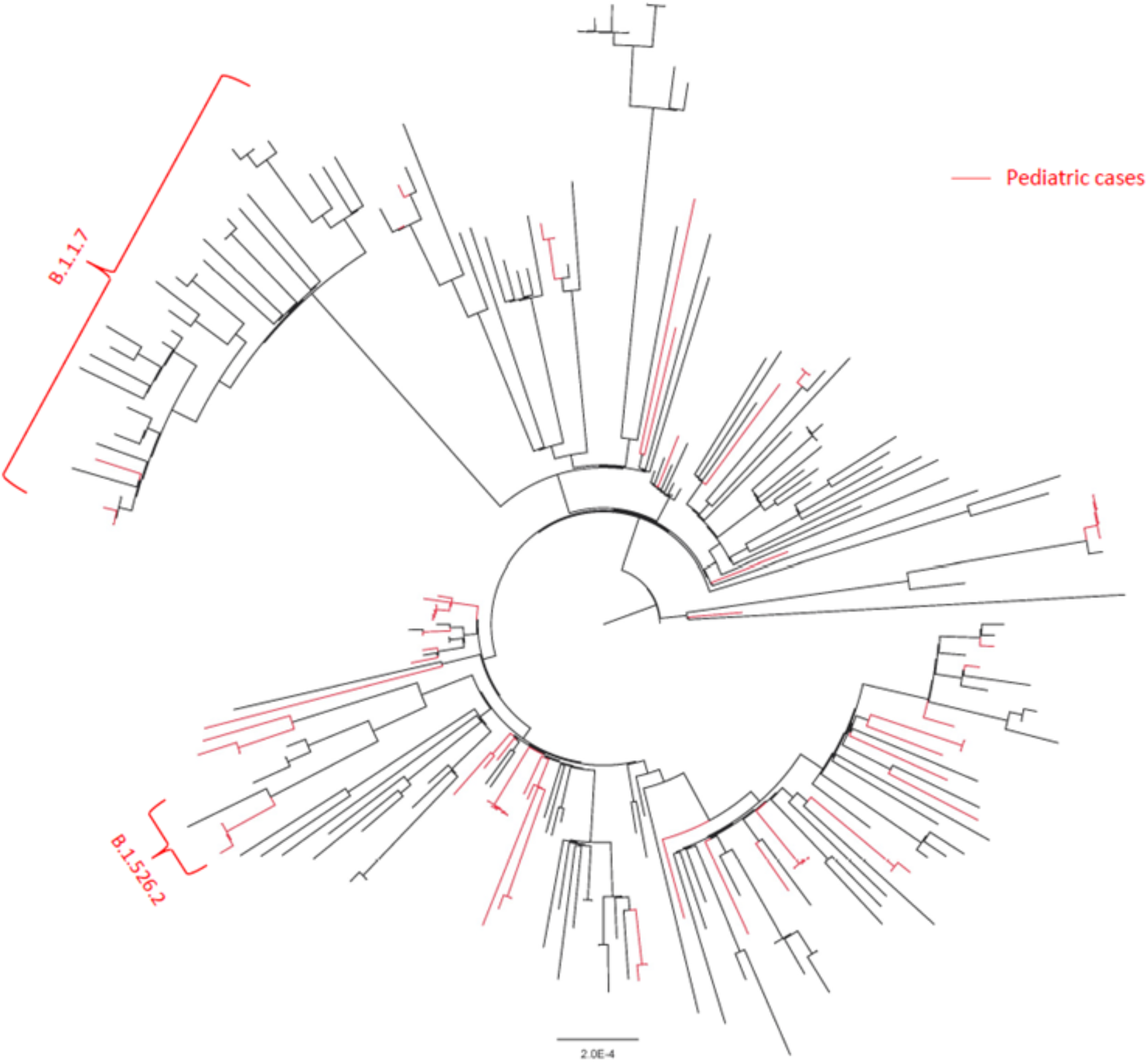
Phylogenetic analysis of pediatric and community SARS-CoV-2 sequences. Maximum likelihood tree generated from pediatric sequences (red) and 183 contemporaneous Massachusetts sequences from GISAID.

## Discussion

As the global COVID-19 pandemic took hold, infected older adults suffered high rates of hospitalization and death while infected children typically experienced paucisymptomatic or asymptomatic infection. While it is now clear that children can become infected with and transmit SARS-CoV-2, viral dynamics in children have been understudied, and a full understanding of the dynamics of infection in children is needed to inform public health policies specific to the pediatric population. Here, we show that pediatric patients of all ages, from infancy to young adulthood, can carry a high SARS-CoV-2 viral load in their upper airways, particularly early in the course of infection, and an elevated viral load corresponds with high levels of viable, replicating virus. Pediatric sequences were largely reflective of those found in the general community and the presence of novel variants was identified.

Our findings have significant implications for both public health policy and the potential role of universal vaccination of pediatric populations in fully curbing the COVID-19 pandemic. As vaccination has rolled out in adult populations, public health policies are being adjusted to account for changes in risk that result from vaccination. Our results emphasize the importance of considering and clarifying how these policy changes relate to children. As adult populations have been vaccinated, pediatric cases have represented a growing proportion of infections, currently accounting for up to 25% of all COVID-19 cases across different regions of the United States[3]. Our results suggest that the low rates of transmission in settings such as schools and daycares cannot be attributed to low viral loads, low rates of viral shedding, or rapid clearance of virus in younger patient populations. As changes in masking and distancing policies are implemented for vaccinated adults, consideration of how and whether policies changes will be applied for children will be critical for ongoing reduction of new COVID-19 cases.

Our results additionally suggest that pediatric populations have the potential to serve as a community reservoir of actively replicating virus, with implications for both new waves of infection and the evolution of viral variants. The duration of natural and vaccine-induced immunity for each vaccine in clinical use are not yet known. If a community reservoir of actively replicating virus is maintained and transmitted within unvaccinated pediatric populations, that population could then serve as a source of new infections as vaccine-induced immunity wanes in vaccinated adult populations. In addition, viral genomic variants were readily identified in the pediatric samples and these variants have the potential to impact viral transmission [23-25], disease severity [26, 27], and vaccine efficacy [28]. Ongoing viral replication within pediatric populations has the potential to serve as a source of existing and new viral variants that interfere with eradication efforts.

Our study has several limitations. First, the data collected here represent a single medical center and affiliated pediatric urgent care/COVID-19 screening clinics. However, these were amongst the few pediatric testing centers encompassing a large catchment area during the duration of this study, and patients enrolled spanned a wide range of symptoms. Additionally, many of these samples were collected early in the pandemic and SARS-CoV-2 variants of interest have shifted over time. Ongoing studies analyzing shifts in virologic features of SARS-CoV-2 infection in children alongside studies of infection in adults are needed to better understand the full reach of the COVID-19 pandemic.

Ultimately, our data suggest that although age is generally protective against severe disease, children, especially early in the infection course, carry high viral loads of SARS-CoV-2, which can include viral variants. Our results underline the importance of defining public health policy with viral dynamics in children in mind and of including pediatric populations in vaccine efforts aimed at eradication.

## Data Availability

Data is available upon request once manuscript is published.

## Acknowledgements

The viral culture work was performed in the Ragon Institute BSL3 core, which is supported in part by the NIH-funded Harvard University Center for AIDS Research (P30 AI060354). This research was supported by the National Heart, Lung, and Blood Institute (5K08HL143183 to LMY), the Department of Pediatrics at Massachusetts General Hospital *for* Children (to LMY), the Massachusetts Consortium for Pathogen Readiness and a gift from Mark and Lisa Schwartz (to JZL).

## Conflicts of Interest

The authors do not report any conflicts of interest.

---

**Supplemental Table 1:** Past medical history of children infected with SARS-CoV-2.

| Past Medical History - Pediatric Patients |
| --- |
| Acute recurrent pancreatitis |
| ADHD |
| Alopecia |
| Asthma |
| Autism |
| Cardiac conduction disorder |
| Chronic kidney disease |
| Coats Plus Syndrome |
| Crohn's disease |
| Cystic Fibrosis |
| Eczema |
| Epilepsy |
| G6PD Deficiency |
| Glutaric acidemia, type 1 |
| Liver transplant |
| Mitochondrial complex 1 deficiency |
| Obesity |
| Obstructive sleep apnea |
| Prematurity |
| Sickle cell trait |
| Speech delay |
| Subglottic stenosis |
| Tracheobronchomalacia |

**Supplemental Table 2:** Demographics, past medical history and disease severity of adults with COVID-19 (n=29) included in analysis of SARS-CoV-2 viral load.

|  | COVID+ adults<br>(n=29) |
| --- | --- |
| Age, average (max, min) | 62 (26,93) |
| Sex, number (%) |  |
| Male | 13 (45) |
| Female | 16 (55) |
| Race, number (%) |  |
| White | 20 (69) |
| Black | 5 (17) |
| Asian | 0 (0) |
| Other | 4 (14) |
| Ethnicity, number (%) |  |
| Hispanic | 4 (14) |
| COVID-19 Severity |  |
| Hospitalized, number (%) | 21 (100) |
| Respiratory support | 22 (78) |
| Outpatient, number (%) | 0 (0) |

**Supplemental Figure 1:**
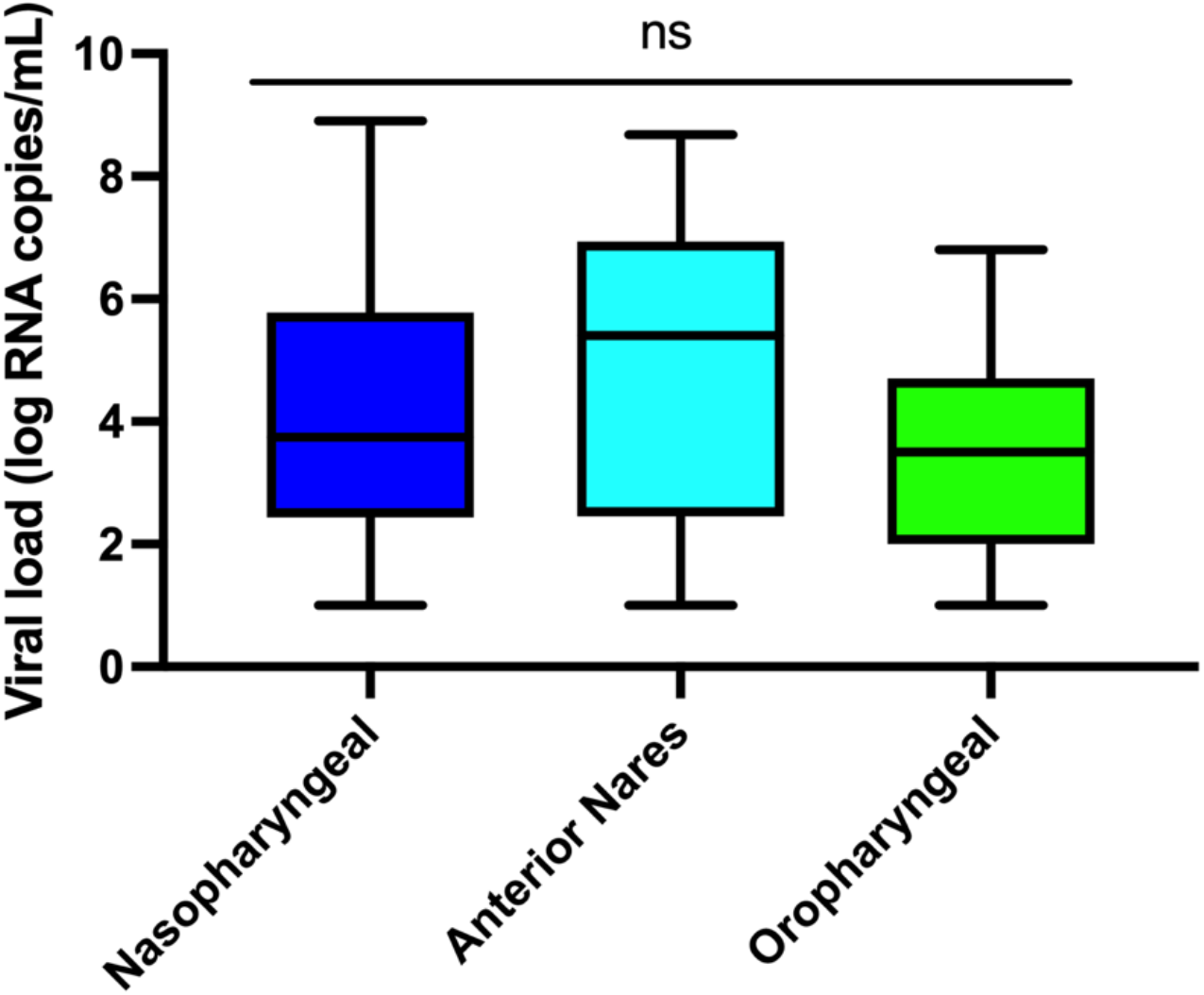
Viral load by sample collection location. Viral load of samples collected from nasopharynx (n=60), anterior nares (n=47), or oropharynx (n=19) were compared and analyzed by ordinary one-way ANOVA. ns = non-significant.

**Supplemental Figure 2:**
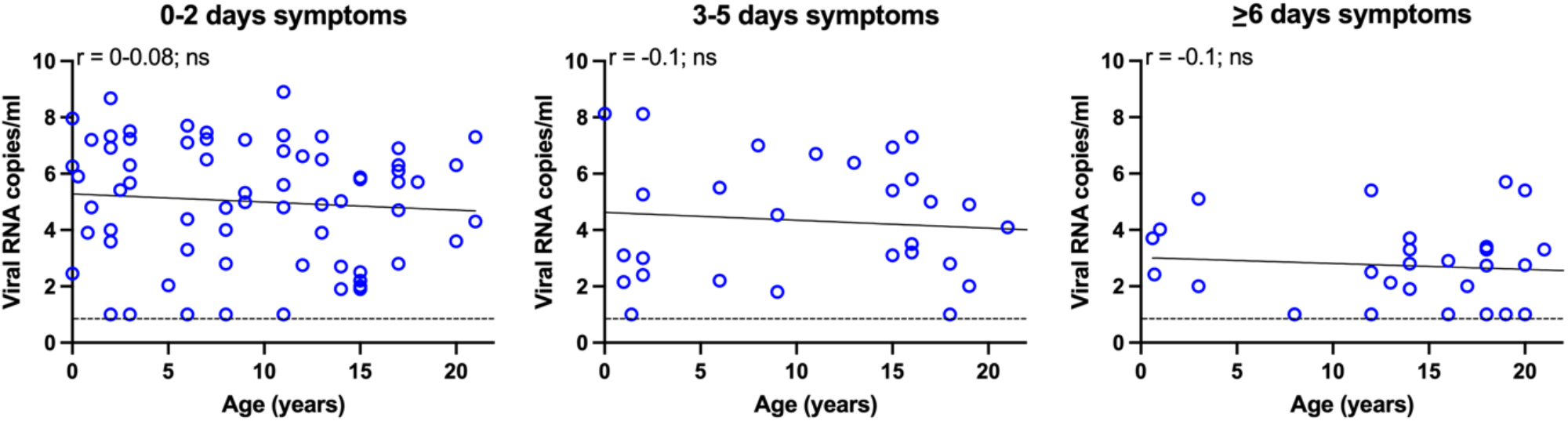
Viral load plotted against pediatric patient age, for patients with 0-2 days symptoms (n=67), 3-5 days symptoms (n=30), and ≥6 days of symptoms (n=29). No significant correlation in any grouping, when analyzed by Pearson correlation. Dotted lines depict limit of detection. ns = not significant.

## Notes

### Competing Interest Statement

The authors have declared no competing interest.

### Author Declarations

Infants, children and adolescents <22 years of age presenting to Massachusetts General Hospital urgent care clinics or the hospital with COVID-19 as determined by PCR testing of nasal specimens (4/2020-4/2021) were offered enrollment in the Institutional Review Board-approved MGH Pediatric COVID-19 Biorepository (IRB # 2020P000955). Nasal samples from adults hospitalized with acute COVID-19 (4/2020-8/2020; enrolled in Institutional Review Board-approved MGH COVID-19 Biorepository, IRB # 2020P000804) were used for comparison of viral load. Demographic and clinical factors were recorded through a combination of manual charts reviews and data extraction from electronic health records (EHR), then collected in a REDCap database [15] through the Partners Electronic Health Record Registry of Pediatric COVID-19 Disease (IRB # 2020P003588).

## References

1. Mehta NS, Mytton OT, Mullins EWS, et al. SARS-CoV-2 (COVID-19): What Do We Know About Children? A Systematic Review. Clin Infect Dis 2020; 71:2469–79.

2. Heald-Sargent T, Muller WJ, Zheng X, Rippe J, Patel AB, Kociolek LK. Age-Related Differences in Nasopharyngeal Severe Acute Respiratory Syndrome Coronavirus 2 (SARS-CoV-2) Levels in Patients With Mild to Moderate Coronavirus Disease 2019 (COVID-19). JAMA Pediatr 2020; 174:902–3.

3. https://services.aap.org/en/pages/2019-novel-coronavirus-covid-19-infections/children-and-covid-19-state-level-data-report/.

4. https://www.mass.gov/info-details/covid-19-response-reporting.

5. Yonker LM, Neilan AM, Bartsch Y, et al. Pediatric Severe Acute Respiratory Syndrome Coronavirus 2 (SARS-CoV-2): Clinical Presentation, Infectivity, and Immune Responses. J Pediatr 2020; 227:45–52 e5.

6. Fialkowski A, Gernez Y, Arya P, Weinacht KG, Kinane TB, Yonker LM. Insight into the pediatric and adult dichotomy of COVID-19: Age-related differences in the immune response to SARS-CoV-2 infection. Pediatr Pulmonol 2020.

7. Soriano-Arandes A, Gatell A, Serrano P, et al. Household SARS-CoV-2 transmission and children: a network prospective study. Clin Infect Dis 2021.

8. L’Huillier AG, Torriani G, Pigny F, Kaiser L, Eckerle I. Culture-Competent SARS-CoV-2 in Nasopharynx of Symptomatic Neonates, Children, and Adolescents. Emerg Infect Dis 2020; 26:2494–7.

9. Weiss A, Jellingso M, Sommer MOA. Spatial and temporal dynamics of SARS-CoV-2 in COVID-19 patients: A systematic review and meta-analysis. EBioMedicine 2020; 58:102916.

10. Fajnzylber J, Regan J, Coxen K, et al. SARS-CoV-2 viral load is associated with increased disease severity and mortality. Nat Commun 2020; 11:5493.

11. Wolfel R, Corman VM, Guggemos W, et al. Virological assessment of hospitalized patients with COVID-2019. Nature 2020; 581:465–9.

12. https://www.cdc.gov/coronavirus/2019-ncov/index.html.

13. Madhi SA, Baillie V, Cutland CL, et al. Efficacy of the ChAdOx1 nCoV-19 Covid-19 Vaccine against the B.1.351 Variant. N Engl J Med 2021.

14. Liu Y, Liu J, Xia H, et al. BNT162b2-Elicited Neutralization against New SARS-CoV-2 Spike Variants. N Engl J Med 2021.

15. Shinde V, Bhikha S, Hoosain Z, et al. Efficacy of NVX-CoV2373 Covid-19 Vaccine against the B.1.351 Variant. N Engl J Med 2021.

16. Lima R, Gootkind EF, De la Flor D, et al. Establishment of a pediatric COVID-19 biorepository: unique considerations and opportunities for studying the impact of the COVID-19 pandemic on children. BMC Med Res Methodol 2020; 20:228.

17. Harris PA, Taylor R, Thielke R, Payne J, Gonzalez N, Conde JG. Research electronic data capture (REDCap)--a metadata-driven methodology and workflow process for providing translational research informatics support. J Biomed Inform 2009; 42:377–81.

18. Choi B, Choudhary MC, Regan J, et al. Persistence and Evolution of SARS-CoV-2 in an Immunocompromised Host. N Engl J Med 2020; 383:2291–3.

19. Gonzalez-Reiche AS, Hernandez MM, Sullivan MJ, et al. Introductions and early spread of SARS-CoV-2 in the New York City area. Science 2020; 369:297–301.

20. Quick J. nCoV-2019 sequencing protocol v1. Available at: https://www.protocols.io/view/ncov-2019-sequencing-protocol-bbmuik6w.

21. Katoh K, Rozewicki J, Yamada KD. MAFFT online service: multiple sequence alignment, interactive sequence choice and visualization. Brief Bioinform 2019; 20:1160–6.

22. Trifinopoulos J, Nguyen LT, von Haeseler A, Minh BQ. W-IQ-TREE: a fast online phylogenetic tool for maximum likelihood analysis. Nucleic Acids Res 2016; 44:W232–5.

23. Zhou B, Thao TTN, Hoffmann D, et al. SARS-CoV-2 spike D614G change enhances replication and transmission. Nature 2021; 592:122–7.

24. Deng X, Garcia-Knight MA, Khalid MM, et al. Transmission, infectivity, and neutralization of a spike L452R SARS-CoV-2 variant. Cell 2021.

25. Davies NG, Abbott S, Barnard RC, et al. Estimated transmissibility and impact of SARS-CoV-2 lineage B.1.1.7 in England. Science 2021; 372.

26. Davies NG, Jarvis CI, Group CC-W, et al. Increased mortality in community-tested cases of SARS-CoV-2 lineage B.1.1.7. Nature 2021; 593:270–4.

27. Funk T, Pharris A, Spiteri G, et al. Characteristics of SARS-CoV-2 variants of concern B.1.1.7, B.1.351 or P.1: data from seven EU/EEA countries, weeks 38/2020 to 10/2021. Euro Surveill 2021; 26.

28. Garcia-Beltran WF, Lam EC, St Denis K, et al. Multiple SARS-CoV-2 variants escape neutralization by vaccine-induced humoral immunity. Cell 2021; 184:2372–83 e9.

